# Scalable federated learning for emergency care using low cost microcomputing: Real-world, privacy preserving development and evaluation of a COVID-19 screening test in UK hospitals

**DOI:** 10.1101/2023.05.05.23289554

**Authors:** Andrew A. S. Soltan, Anshul Thakur, Jenny Yang, Anoop Chauhan, Leon G. D’Cruz, Phillip Dickson, Marina A. Soltan, David R. Thickett, David W. Eyre, Tingting Zhu, David A. Clifton

**Affiliations:** Oxford University Hospitals NHS Foundation Trust; RDM Division of Cardiovascular Medicine, University of Oxford; Institute of Biomedical Engineering, Dept. Engineering Science, University of Oxford; Portsmouth Hospitals University NHS Foundation Trust; Bedfordshire Hospitals NHS Foundation Trust; The Queen Elizabeth Hospital, University Hospitals Birmingham NHS Foundation Trust; Institute of Inflammation and Ageing, University of Birmingham; Big Data Institute, Nuffield Department of Population Health, University of Oxford; NIHR Health Protection Research Unit in Healthcare Associated Infections and Antimicrobial Resistance at University of Oxford in partnership with Public Health England; NIHR Oxford Biomedical Research Centre

## Abstract

**Background:** Tackling biases in medical artificial intelligence requires multi-centre collaboration, however, ethical, legal and entrustment considerations may restrict providers’ ability to participate. Federated learning (FL) may eliminate the need for data sharing by allowing algorithm development across multiple hospitals without data transfer.

Previously, we have shown an AI-driven screening solution for COVID-19 in emergency departments using clinical data routinely available within 1h of arrival to hospital (vital signs & blood tests; CURIAL-Lab). Here, we aimed to extend and federate our COVID-19 screening test, demonstrating development and evaluation of a rapidly scalable and user-friendly FL solution across 4 UK hospital groups.

**Methods:** We supplied a Raspberry Pi 4 Model B device, preloaded with our end-to-end FL pipeline, to 4 NHS hospital groups or their locally-linked research university (Oxford University Hospitals/University of Oxford (OUH), University Hospitals Birmingham/University of Birmingham (UHB), Bedfordshire Hospitals (BH) and Portsmouth Hospitals University (PUH) NHS trusts). OUH, PUH and UHB participated in federated training and calibration, training a deep neural network (DNN) and logistic regressor to predict COVID-19 status using clinical data for pre-pandemic (COVID-19-negative) admissions and COVID-19-positive cases from the first wave. We performed federated prospective evaluation at PUH & OUH, and external evaluation at BH, evaluating the resultant global and site-tuned models for admissions to the respective sites during the second pandemic wave. Removable microSD storage was destroyed on study completion.

**Findings:** Routinely collected clinical data from a total 130,941 patients (1,772 COVID-19 positive) across three hospital groups were included in federated training. OUH, PUH and BH participated in prospective federated evaluation, with sets comprising 32,986 patient admissions (3,549 positive) during the second pandemic wave. Federated training improved DNN performance by a mean of 27.6% in terms of AUROC when compared to models trained locally, from AUROC of 0.574 & 0.622 at OUH & PUH to 0.872 & 0.876 for the federated global model. Performance improvement was more modest for a logistic regressor with a mean AUROC increase of 13.9%. During federated external evaluation at BH, the global DNN model achieved an AUROC of 0.917 (0.893-0.942), with 89.7% sensitivity (83.6-93.6) and 76.7% specificity (73.9-79.1). Site-personalisation of the global model did not give a significant improvement in overall performance (AUROC improvement <0.01), suggesting high generalisability.

**Interpretations:** We present a rapidly scalable hardware and software FL solution, developing a COVID-19 screening test across four UK hospital groups using inexpensive micro-computing hardware. Federation improved model performance and generalisability, and shows promise as an enabling technology for deep learning in healthcare.

Funding University of Oxford Medical & Life Sciences Translational Fund/Wellcome

## Evidence before this study

International consortia have highlighted the importance of adequate representation in health AI datasets, with multiple reviews identifying shortfalls in diversity most commonly due to a lack of systematic data-sharing. Dame Fiona Caldicott’s 2013 report set out the governance challenges facing healthcare providers participating in data-sharing, and the recent emergence of federated learning (FL) has been highlighted as a promising solution for providers to participate in medical AI development. We searched PubMed for applications of FL in hospitals (search terms: “federated learning” AND (“hospital” OR “hospitals#x201D;) AND (#x201C;screen” OR “screening” OR “diagnosis” OR “prognosis” OR “prognostication” OR “outcomes#x201D;)), finding 32 results to November 01, 2022, of which 5 describe implementations of FL in secondary care using medical imaging (chest x-ray and computerised tomography) for diagnosis and prognostication in COVID-19. To our knowledge, no works to-date describe the use of micro-computing alongside FL to assist in its deployment within hospitals, or demonstrated FL-driven screening using routinely collected vital signs and blood tests which are much more available and do not require use of ionising radiation.

### Added value of this study

Here we present a development, validation and deployment of a Federated Learning solution across four UK hospital groups, extending our prior work on AI-driven screening for COVID-19 in emergency care. To our knowledge, our study is the first to couple an FL pipeline with deployment of micro-computing hardware in a real-world secondary care setting. We select the commercially produced Raspberry Pi model 4B for its low cost ($45-80), thereby enabling rapid scale, and removable micro-SD card storage which is securely destroyed on completion of participation to prevent subsequent data loss. Our results show a large improvement in performance on federation of the model, which is more marked for deep learning than a traditional statistical method, and robust and generalisable performance across 3 hospital groups evaluating on prospective cohorts. Our study is the largest secondary-care FL study to date by number of patient encounters, including the routinely collected clinical data for over 160,000 participants attending 4 hospital groups that serve a combined population of 3.5 million.

### Implications of all the available evidence

Our study offers a paradigm for future FL research within secondary care settings, enabling AI models to be developed and validated in the real-world without transfer of patient data. Federated learning may be an enabling technology for deep learning, and micro-computing hardware may have a role in implementation.

### Background

Legal, ethical and entrustment challenges surround use of patient data for artificial intelligence (AI) research, with mounting public concern regarding unintended use, misuse and reidentification attacks^1–4^. Concerns around organisational ability to maintain control of data once transferred off-premises, and fear of potential consequences, were identified by the Caldicott review as drivers for unduly restrictive information governance rules, reduced co-operation, and a ‘culture of anxiety’^5^. These considerations may hamper efforts to improve diversity within training sets^4, 6^.

Client-server federated learning (FL) has emerged as a leading privacy enhancing technology (PET) for collaborative development of AI models without transfer of data outside of participating organisations^7, 8^. Whereas in classical machine learning the training process would take place on a central server where data is aggregated, in FL data remains under the custody of the supplying organisation and training/evaluation processes occur locally. Following each round of local training, model weights -and not patient data-are transferred from clients to a central server where aggregation is performed to create a ‘global model’, and is re-circulated to clients for iteration^9, 10^.

FL may encourage healthcare providers to participate in AI research, thereby reducing development time, improving representation and facilitating international collaboration^6, 11^. Successful implementations have included prediction of mechanical ventilation/death in COVID-19 across 20 hospitals using NVIDIA’s Clara Platform [California, USA]^8^. However, to date real-world implementations of FL in hospital settings have been limited in number^12–16^, and some approaches advocate use of an intermediate platform^13^. Experimental works have shown some promising results for federated COVID-19 screening using medical imaging (chest x-ray & computerised tomography) however, these studies have been within a simulated setting without a real-world deployment, and with modest sample sizes^17, 18^. Moreover, complexity of user interface and set-up has been identified as a barrier to adoption of health-AI^19^, and may also limit the uptake of FL amongst providers.

An ‘Internet of Medical Things’ (IoMT), in which connected micro-computing devices are used to deliver care, has shown promise for improving engagement, outcomes and cost-effectiveness^20, 21^. Successful IoMT applications have included patient-facing wearables^22–24^, however limited work has explored applications of micro-computing within healthcare providers or a deployment of health-AI in secondary care.

Our group has previously developed, validated and piloted an AI screening test for COVID-19, for use in emergency departments (ED)^25, 26^. The CURIAL-Lab test aims to reduce nosocomial transmission and ease operational pressures by utilising clinical data routinely collected within 1 h of a patient arriving in hospital (vital signs, full blood count, liver function tests, urea & electrolytes, and C-reactive protein) to provide a high confidence result-of-exclusion. The initial work, highlighted in a 2022 editorial^27^, included design considerations to prioritise patient confidentiality when working across multiple hospital groups. We asked NHS trusts to de-identify patient data at source and employed secure protocols for transfer to a trusted server at the University of Oxford where analysis was performed. However, de-identification processes can lead to a loss of informative predictors^28^, and may alone be insufficient to safeguard privacy in the event of a data leak^29^.

To eliminate the need for transfer of patient data, we propose and deploy a user-friendly federated training, calibration and evaluation pipeline for COVID-19 screening across four UK hospital groups (CURIAL-Fed-Lab). We combine a custom software pipeline with micro-computing hardware to provide an end-to-end solution, supplying each participating hospital group or their linked research University with a pre-configured Raspberry Pi 4B [£40-85, Raspberry Pi Ltd, Cambridge, UK] running the commercially supported Ubuntu Desktop operating system. Strengths of our approach include its ease-of-use (Supplementary Figure S1), eliminating the need for local technical expertise, and use of inexpensive micro-computing devices to permit rapid scale. Further, our solution uses removable microSD storage which can be destroyed on completion to prevent subsequent data loss^30^.

## Methods

### Privacy-preserving federated learning for COVID-19 screening in ED

Four NHS hospital groups participated in the CURIAL-Fed-Lab study (Figure 1); these included Oxford University Hospitals NHS Foundation Trust (OUH), Portsmouth Hospitals University Trust (PUH), University Hospitals Birmingham NHS Foundation Trust (UHB) and Bedfordshire Hospitals NHS Foundation Trust (BH). Further details are provided in **Appendix A**. OUH, UHB and PUH participated in federated training and calibration. OUH, PUH and BH participated in federated evaluation. Data extracted from BH was additionally used for a centralised evaluation.

**Figure 1:**
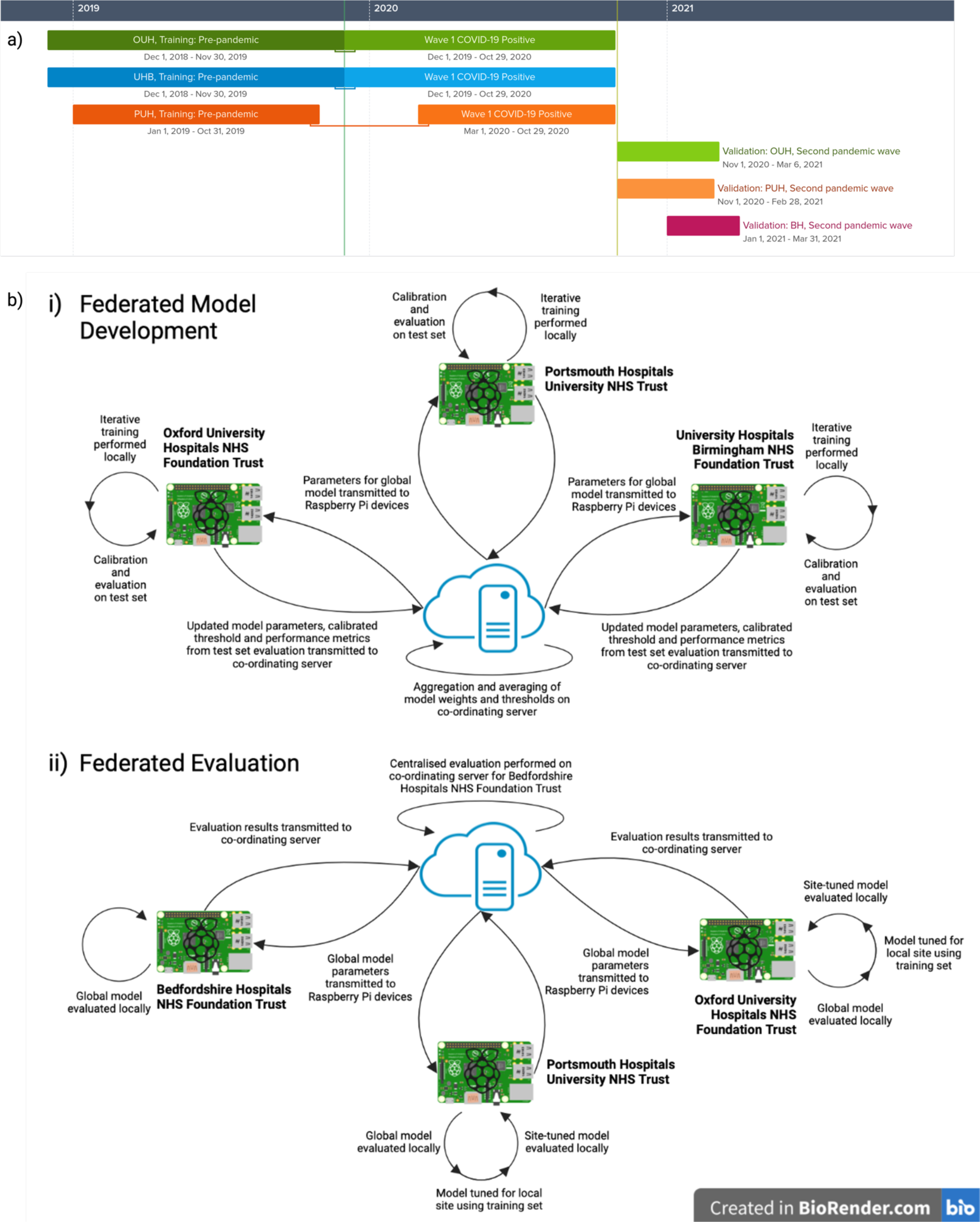
Overview of study design. (a) Timeline showing derivation of training and prospective evaluation cohorts. (b) Federated training & evaluation study design. bi) Deidentified patient data is extracted by NHS trusts and loaded on to Raspberry Pi devices held locally within the hospital group or its linked research university. Machine learning models are trained locally and calibrated and evaluated on a locally-held test set. Model weights, thresholds and evaluation results are transmitted to a co-ordinating server, where aggregation and averaging is performed to form a global model. Updated weights for the new global model are transmitted to local devices, facilitating the next round of training. 150 rounds are performed. bii) Following each training of round, weights for the trained global model are transmitted to the devices at local participating sites. Federated evaluation is performed by applying the models to prospective cohorts of patients admitted to hospital during the second wave of the UK COVID-19 Pandemic at OUH, PUH and BH. For sites also contributing to training (OUH & PUH), an additional step of site-personalisation is performed and the personalised model evaluated. Evaluation results are transmitted to the co-ordinating server for reporting. For quality assurance, centralised evaluation is also performed on the co-ordinating server for BH.

**Table 1:** Clinical predictors within the CURIAL-Fed-Lab model.

| Feature Set | Constituents |
| --- | --- |
| Vital Signs | Heart rate, respiratory rate, oxygen saturations, blood pressure, temperature, oxygen delivery device level |
| Full Blood Count (FBC) | Haemoglobin, haematocrit, mean cell volume, white cell count, neutrophil count, lymphocyte count, monocyte count, eosinophil count, basophil count, platelets |
| Urea & Electrolytes (U&Es) | Sodium, potassium, creatinine, urea, eGFR |
| Liver Function (LF) Tests & CRP | Albumin, alkaline phosphatase, ALT, bilirubin, C-Reactive Protein |

### Implementation

We performed client-server based federated training & evaluation, supplying a Raspberry Pi 4 Model B device (the client; Raspberry Pi Limited, Cambridge, UK) configured with at least 2Gb of Random Access Memory (RAM) and 32Gb removal microSD storage, to participating NHS hospital trusts or their linked research university. We pre-installed Ubuntu 22.04.1 LTS, necessary dependency packages (see Appendix A), and our custom FL pipeline based on the Flower framework (code available via Github and as a flashable disk image for the Raspberry Pi)^31^. We selected the Raspberry Pi 4 Model B as a commercially available and inexpensive device (£45-85), thereby allowing for rapid scale, and for its removable microSD storage enabling participating Trusts to securely destroy media containing the patient data extract on completion. No patient data was transmitted from clients to co-ordinating server as part of the federated training, thereby preserving privacy (see Appendix A). Source code was available to participating sites for review. Clients were operated on-premises by the respective NHS Trusts at PUH and BH, and by the locally-linked University within a shared research network at OUH and UHB (University of Oxford and University of Birmingham respectively, within the NIHR Biomedical Research campuses). Where necessary, firewall rules were instated to permit two-way communication between client and server through a pre-agreed port (see Appendix A for further details). On completion of participation, participating sites were directed to remove and securely dispose of the microSD card following organisational procedures for hardware disposal.

### Study Populations for Federated Training & Evaluation

We provided participating NHS Trusts with inclusion and exclusion criteria for extraction from Electronic Healthcare Records (EHR), alongside requested clinical parameters (**Supplementary Table S1**). Screening against criteria, followed by de-identification and extraction, was performed by each participating NHS Trust and enforced programmatically within the analysis pipeline. For both training & evaluation, patients included had an unscheduled acute or emergency care admission, received a blood draw on arrival, and were aged over 18. Patients who had opted out of EHR research or who did not receive routine laboratory blood tests within 24h of arriving at hospital were excluded.

Due to incomplete penetrance of testing and imperfect PCR sensitivity during the first pandemic wave, there is uncertainty in the viral status of patients presenting during the early pandemic who were untested or tested negative. Therefore as previously, for training we selected a pre-pandemic control cohort (attending hospital prior to December 1, 2019) to ensure absence of disease in patients labelled as COVID-19-negative. Patients presenting during the first wave, between December 1, 2019 and October 29, 2020, with PCR confirmed SARS-CoV-2 infection formed the COVID-19-positive (cases) training cohort. For federated evaluation, we selected independent prospective sets of adult patients admitted to OUH, PUH and BH during the second pandemic wave in the UK, defined as after November 1^st^, 2020. Evaluation included patients receiving confirmatory molecular testing with either a positive or negative result, excluding indeterminate or invalid results. Further information on training & evaluation cohorts, alongside confirmatory testing method, are provided in **Appendix A**.

Clinical features extracted for each presentation included demographics (age, gender, ethnicity), results of first-performed blood testing, blood gases, vital signs measurements and results of molecular testing for SARS-CoV-2 (**Supplementary Table S1**). As previously, we selected routine blood tests to include the full blood count (FBC), urea and electrolytes (U&E), liver function tests (LFT), and C-reactive protein (CRP) because they are widely performed within existing care pathways in emergency departments and results are typically available within 1 h^25^. Staff at participating organisations were directed to load the data extracts on to the supplied Raspberry Pi device and activate the study application (Appendix A & Supplementary Figure S1).

### Federated pipeline

We deployed a custom analysis pipeline, pre-installed on the Raspberry Pi 4B devices, to locally i) standardise the anonymised data extracts in to a common format, ii) perform normalisation and imputation, iii) federated training, and iv) federated evaluation. Feature names, result representations and units, and SARS-CoV-2 PCR results were programmatically standardised into a common format on-device (Appendix A). To ensure accurate cohort eligibility, inclusion & exclusion criteria were programmatically re-enforced. Missing data were imputed by selecting the median value of the training population, as we previously showed stability of model performance across multiple imputation strategies (mean, median and age-based mean)^26^. Training population median values for each site were autonomously transmitted to the federated server, to facilitate imputation for sites performing evaluation only. We performed normalisation by scaling training data to a range between 0 and 1, aiming to mitigate against biases towards features with large numerical values. As previously, patients with PCR-confirmed SARS-CoV-2 infection during the first wave were matched with pre-pandemic controls across three demographic factors (ethnicity, gender and age to within +/-4 years per participant). A case-control ratio of 1:10 was selected during training to limit the degree of class imbalance. 20% of the training set was reserved for internal evaluation and calibration.

### Federated Training

We performed 150 rounds of federated training across three contributing hospital groups (OUH, PUH, UHB; Fig 1), implementing the FedAvg algorithm^10^. Initial model parameters were randomly generated and clients trained a local model on their individual training sets. Following local training, local models were evaluated and model parameters transmitted by clients to the central server for aggregation and calculation of a global model. The new global model parameters were subsequently transmitted to the clients, replacing the locally-held model, prior to the next training round. To maximise data utilisation, we sampled each participating site (client) for every round of training. Locally-held datasets were not accessible to the server during training.

We performed federated training for two different binary classifiers aiming to predict COVID-19 PCR result. First, as a base case, we trained a Logistic regression (LR) classifier with an L2 ridge regression regularisation penalty, performing 5 iterations over the training data per round. Next, we trained a deep neural network (DNN) comprising of an input layer, a dense hidden layer with 10 nodes, a dropout regularisation layer (rate 0.5) to mitigate overfitting, and an output layer. The rectified linear unit (ReLU) activation function was used for the hidden layers and the sigmoid activation function in the output layer. For updating model weights, the Adaptive Moment Estimation (Adam) optimizer was used with a learning rate of 0.0001. For initial local training and each subsequent round of FL, we configured the clients to iterate over the training data for up to 50 epochs with early stopping if the AUC on the held-out test set did not improve over 15 sequential epochs. Each client tracked performance of its best-performing local model when evaluated on the held-out test set after each epoch, transmitting weights for this best model to the server for aggregation and updating of the global model.

### Testing and calibration

Following each round of federated training, local models were calibrated by selecting the prediction threshold required to achieve a sensitivity of 85% on the held-out test. Evaluation results for the test set, and the selected threshold, were transmitted to the co-ordinating server for aggregation.

### Federated Evaluation of Global Model

We performed prospective federated evaluation by locally validating our global model, calibrated to 85% sensitivity, for emergency hospital admissions during the second wave of the COVID-19 pandemic at OUH, PUH, and BH. Model predictions were evaluated by comparison to results of confirmatory molecular testing performed on admission (SARS-CoV-2 laboratory PCR and the point-of-care PCR devices SAMBA-II and Panther; Appendix A).

For sites both contributing to federated training and evaluation (OUH & PUH), calibration was performed by selecting the locally-determined threshold identified during calibration on the held-out test set. Missing data were imputed using median values of the training population at the local site. For sites performing federated evaluation only (BH), we selected the threshold by performing autonomous server-side aggregation and averaging (mean) of the optimum local thresholds at each of the three sites contributing to training (OUH, PUH and UHB). Missing data at BH were imputed by autonomously calculating the mean of the median population values for the three contributing sites on the evaluation server and transmitting the result to the BH client, eliminating the need for any transfer or aggregation of patient data between sites. Summary statistical measures of the results of federated evaluation (sensitivity, specificity, predictive values and AUROC) were transmitted to the server for reporting.

### Site-specific model tuning

To investigate model sensitivity to distribution shifts between contributing NHS Trusts, thereby assessing generalisability, we investigated whether performing local tuning of the global model would affect performance during evaluation. Following each round of training, we tuned the global model by locally performing a final cycle of training on the local training set for sites contributing to training (OUH & PUH). Model performance was assessed on the prospective validation set and compared with the untuned global model.

### Centralised (server-side) Evaluation

To verify integrity of the federated evaluation, we additionally performed centralised evaluation by validating the global model for all patients admitted to BH after each training round on the central server. The BH data extract was transferred to the server to facilitate this. For centralised evaluation, the mean threshold across all sites contributing to training was used. Median population values from each training site were transmitted to the server, and a mean of the median values used to impute missing data. To understand the impacts of individual features on model predictions, we calculated SHAP (SHapley Additive exPlanations) values for the global models using a subset of 400 cases^32^.

### Statistical Analysis Methods

Model performance was evaluated during testing and prospective evaluation in terms of area under receiver operating characteristic curves (AUROC), sensitivity, specificity, positive predictive value (PPV), negative predictive value (NPV), and F1 score. We compared the performance of (i) locally-trained models with federated global models, (ii) federated global models with site-tuned variations, and (iii) the global LR model with the global DNN model, within the federated pipeline, using DeLong’s Test^33^.

### Ethics

NHS Health Research Authority (HRA) approval (IRAS ID 281832).

## Results

### Study Populations

Three NHS trusts (OUH, UHB and PUH) participated in federated training, contributing routinely collected clinical data from 129,169 patients admitted to hospital prior to the pandemic and 1,772 patients admitted with PCR-confirmed COVID-19. OUH, PUH and BH participated in prospective federated evaluation, comprising 32,986 patients admitted during the second pandemic wave, of whom 3,549 tested positive. During the evaluation period, prevalence was similar between PUH and BH (11.2% & 12.3%, Fisher’s Exact p=0.29), but lower at OUH (10.3%; p=0.01 for PUH & p=0.04 for BH). Patients admitted to OUH and BH had similar ages (67 years, IQR 31, for OUH versus 68 years, 34, for BH; Kruskal Wallis p=0.31), whereas patients admitted to PUH were older (69 years, 34; p<0.0001).

**Table 2:** Summary population characteristics for (a) training cohorts at OUH, UHB and PUH, divided by pre-pandemic control patients and COVID-19-cases during the first wave of the UK COVID-19 pandemic, (b) prospective validation cohorts of patients admitted to OUH, PUH & BH during the second wave of the UK COVID-19 epidemic. * indicates merging for statistical disclosure control.

|  | a) Training cohorts: Pre-pandemic & Wave 1 COVID-19 positive |  |  |  |  |  | b) Wave 2: Prospective evaluation cohorts |  |  |
| --- | --- | --- | --- | --- | --- | --- | --- | --- | --- |
|  | Oxford University Hospitals |  | University Hospitals Birmingham NHS Foundation Trust |  | Portsmouth Hospitals University NHS Trust |  | Oxford University Hospitals | Portsmouth Hospitals University NHS Trust | Bedfordshire Hospitals NHS Foundation Trust |
| Cohort | Pre-pandemic: December 1, 2018 - November 30, 2019 | Wave 1, COVID19+: December 1, 2019 - October 29, 2020 | Pre-pandemic: December 01, 2018 - November 30, 2019 | Wave 1, COVID19+: December 01, 2019 - October 29, 2020 | Pre-pandemic: January 01, 2019 - October 31, 2019 | Wave 1, COVID19+: March 1, 2020 - October 29, 2020 | Wave 2: November 01, 2020 - March 06, 2021 | Wave 2: November 01, 2020 - February 28, 2021 | Wave 2: January 1, 2021 - March 31, 2021 |
| n, patients | 68,496 | 816 | 12,901 | 439 | 47,772 | 517 | 18,543 | 13,260 | 1183 |
| n, COVID-19 genome test +ve (%) |  | 816 |  | 439 |  | 517 | 1,916 (10.3%) | 1,488 (11.2%) | 145 (12.3%) |
| Sex: |  |  |  |  |  |  |  |  |  |
| - Male (%) | 32,286 (47.14) | 435 (53.31) | 5,900 (45.73) | 257 (58.54) | 20,345 (42.59) | 315 (60.93) | 9,235 (49.8) | 5,816 (43.86) | 629 (53.17) |
| - Female (%) | 36,210 (52.86) | 381 (46.69) | 7,001 (54.27) | 182 (41.46) | 27,425 (57.41) | 202 (39.07) | 9,308 (50.2) | 7,442 (56.12) | 553 (46.75) |
| Age, yr (IQR) | 64.0 (44.0-79.0) | 69.0 (54.0-81.0) | 61.0 (40.0-79.0) | 65.0 (51.0-81.0) | 65.0 (41.0-79.25) | 73.0 (60.0-83.0) | 67.0 (49.0-80.0) | 69.0 (48.0-82.0) | 68.0 (48.0-82.0) |
| Ethnicity: |  |  |  |  |  |  |  |  |  |
| -White (%) | 56,295 (82.19) | 554 (67.89) | 8,486 (65.78) | 228 (51.94) | 37,321 (78.12) | 367 (70.99) | 14,079 (75.93) | 9,954 (75.07) | 1,030 (87.07) |
| -Not Stated (%) | 8,050 (11.75) | 149 (18.26) | 1,231 (9.54) | 69 (15.72) | 9,355 (19.58) | 131 (25.34) | 3,340 (18.01) | 3,014 (22.73) | * |
| -South Asian (%) | 1,507 (2.2) | 34 (4.17) | 1867 (14.47) | 96 (21.87) | 246 (0.51) | * | 369 (1.99) | 62 (0.47) | 71 (6.0) |
| -Chinese (%) | 145 (0.21) | * | 60 (0.47) | * | 39 (0.08) | * | 44 (0.24) | 14 (0.11) | * |
| -Black (%) | 813 (1.19) | 28 (3.43) | 666 (5.16) | 21 (4.78) | 229 (0.48) | * | 238 (1.28) | 72 (0.54) | 36 (3.04) |
| -Other (%) | 1,112 (1.62) | 39 (4.78)* | 347 (2.69) | 25 (5.69)* | 358 (0.75) | 19 (3.68)* | 347 (1.87) | 94 (0.71) | 33 (2.79) * |
| -Mixed (%) | 574 (0.84) | 12 (1.47) | 244 (1.89) | * | 212 (0.44) | * | 126 (0.68) | 50 (0.38) | 13 (1.1) |

To assess the effect of federation on model performance during development, we evaluated the global model on the held-out test set after each round of training (**Supplementary Figure S1**). Federation improved classifier stability for LR, achieving optimum performance at all sites within 10 rounds. The DNN classifier demonstrated sustained improvement in AUROC across sequential rounds, with plateauing improvement after approximately 50 rounds.

### Federated Prospective (OUH, PUH) & External (BH) Evaluation

We compared the trained local models, trained using a maximum 50 epochs, with the final federated global and site-personalised models by evaluation on the prospective validation sets at sites participating in both training & evaluation (Figure 2). Federated training significantly improved AUROC of the Logistic Regressor from 0.685 (95% CIs: 0.673-0.698) for the locally-trained model to 0.829 (0.819 −0.839) for the global model at OUH, and from 0.731 (0.718 - 744) to 0.865 (0.854 - 0.876) at PUH (DeLong p < 0.0001 for both), representing a mean 13.9% increase in AUROC. The performance improvement due to federation was more marked for the DNN model, improving from 0.574 (0.560 - 0.589) to 0.872 (0.862 - 0.882) at OUH, and 0.622 (0.608 - 0.637) to 0.876 (0.865 - 0.886) at PUH (p < 0.0001 for both), a mean 27.6% increase in AUROC; possibly reflecting the high data requirements of deep neural networks.

**Figure 2:**
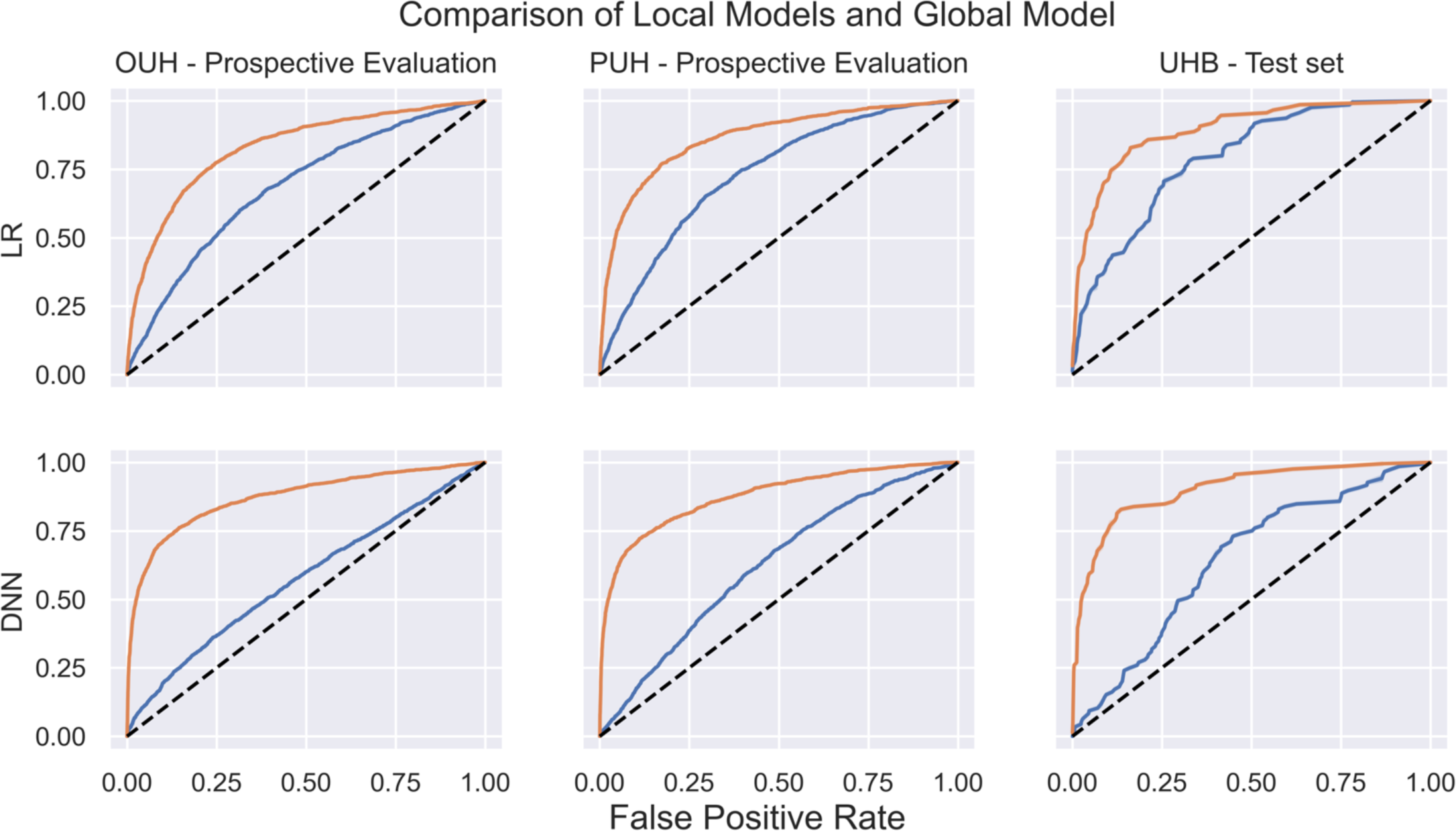
Receiver operating characteristic curves showing performance of (i) locally trained models prior to federation (blue), and (ii) the federated global model (orange) during prospective validation at OUH & PUH, and during evaluation on the locally-held test set at UHB. The area between receiver operating characteristic curves denotes the performance improvement on federation.

When the final global model was externally and prospectively evaluated for all patients admitted to Bedfordshire Hospitals NHS Foundation Trust between January 1, 2021 and March 31, 2021, both LR and DNN global models demonstrated high classification performance (AUROC: 0.878, 95% CI 0.851-0.904, for the LR Global model, and 0.917, 0.893-0.942, for the DNN Global Model). Federated calibration was effective, achieving sensitivities of 83.4% and 89.7% for the LR and DNN respectively during external evaluation. Both global models showed stable performance across the three evaluating sites (AUROCs range 0.829 to 0.878, 95% CIs range 0.819-0.904, for LR, and 0.872 to 0.917, 95% CIs range 0.862-0.942, for DNN; Table 3). As during training, the improvement in validation performance brought about by federation was more marked for DNN than LR, achieving plateau after approximately 75-100 rounds of federation when compared to approximately 10 for LR (Figure 3). Although the global DNN model outperformed the global LR model at BH (DeLong p=0.0011) and OUH (p<0.0001), it performed similarly at PUH (p=0.81).

**Figure 3:**
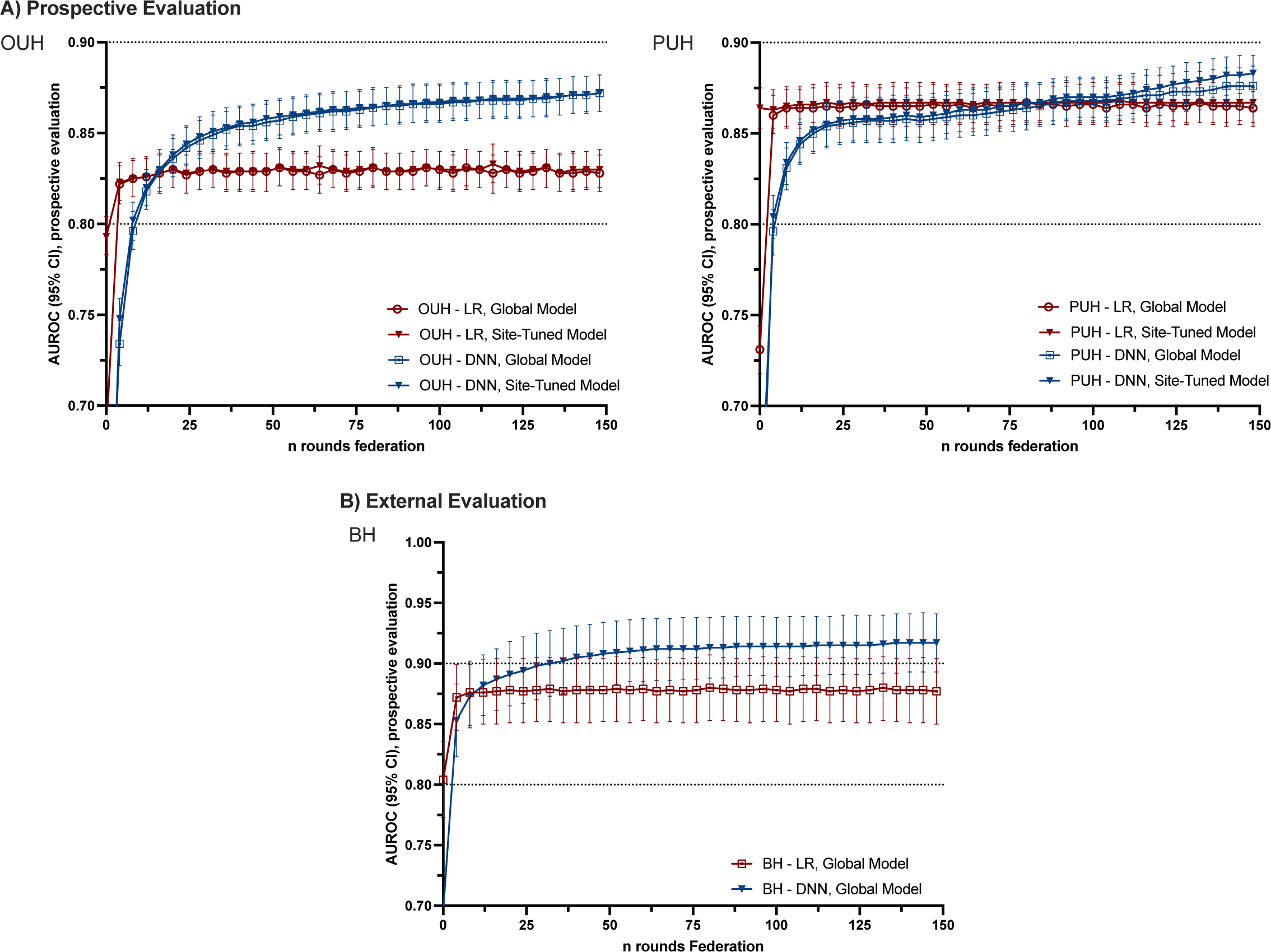
Effect of increasing rounds of federated training on performance of LR & DNN models (AUROC +/-95% CIs) during federated evaluation. (a) Prospective evaluation of both global and site-tuned models for patients admitted to OUH & PUH.

**Figure 4:**
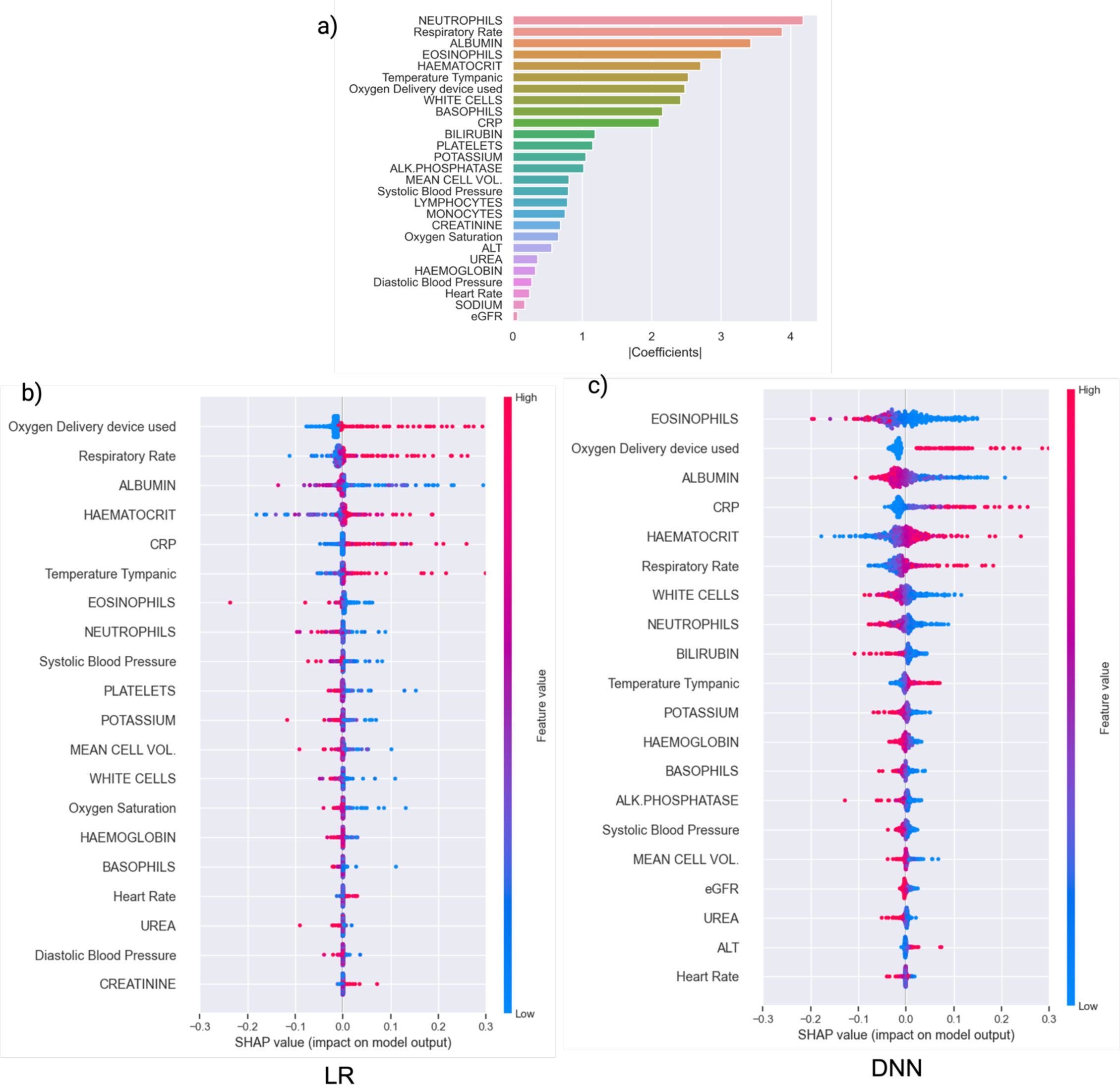
Explainability analyses. (a) logistic regression coefficient scalars within the final global model. (b) and (c) SHAP values for the 20 features with greatest impact on predictions made by the LR and DNN global models respectively, calculated during centralised external validation at BH and shown as beeswarm plots. Each dot represents a patient attending BH during the prospective evaluation period. Positive SHAP values indicate a change in the expected model prediction towards testing positive for COVID-19. Features are shown in descending order of mean absolute SHAP value, with most impactful features shown at the top.

**Table 3:** Performance of calibrated local and federated (global & site-personalised) models for identifying patients being admitted to hospital with COVID-19 during prospective evaluation. AUROC, Sensitivity, specificity, and predictive values are reported alongside 95% CIs. Calibration was performed locally during training for sites participating in both federated training and evaluation (OUH and PUH), and was federated for sites participating only for evaluation (BH).

| Model | AUROC | Sensitivity | Specificity | Accuracy | PPV | NPV | F1 |
| --- | --- | --- | --- | --- | --- | --- | --- |
| **Oxford University Hospitals NHS Foundation Trust** |  |  |  |  |  |  |  |
| LR: Local Model | 0.685 (0.673 - 0.698) | 86.8% (85.3 - 88.3) | 32.7% (32.0 - 33.4) | 38.3% (37.6 - 39.0) | 12.9% (12.4 - 13.5) | 95.6% (95.0 - 96.1) | 0.225 |
| LR: Federated Global Model | 0.829 (0.819 - 0.839) | 81.1% (79.3 - 82.8) | 70.1% (69.4 - 70.8) | 71.2% (70.6 - 71.9) | 23.8% (22.8 - 24.9) | 97.0% (96.7 - 97.3) | 0.368 |
| LR: Federated Site-Personalised Model | 0.83 (0.819 - 0.84) | 80.0% (78.1 - 81.7) | 71.4% (70.7 - 72.1) | 72.3% (71.6 - 72.9) | 24.4% (23.3 - 25.5) | 96.9% (96.5 - 97.2) | 0.374 |
| DNN: Local Model | 0.574 (0.56 - 0.589) | 83.4% (81.6 - 85.0) | 20.6% (20.0 - 21.2) | 27.1% (26.5 - 27.7) | 10.8% (10.3 - 11.3) | 91.5% (90.6 - 92.3) | 0.191 |
| DNN: Federated Global Model | 0.872 (0.862 - 0.882) | 80.8% (79.0 - 82.5) | 78.6% (78.0 - 79.3) | 78.9% (78.3 - 79.5) | 30.4% (29.1 - 31.7) | 97.3% (97.0 - 97.5) | 0.442 |
| DNN: Federated Site-Personalised Model | 0.873 (0.863 - 0.883) | 81.1% (79.2 - 82.7) | 78.0% (77.3 - 78.6) | 78.3% (77.7 - 78.9) | 29.8% (28.5 - 31.0) | 97.3% (97.0 - 97.5) | 0.435 |
| **Portsmouth Hospitals University NHS Trust** |  |  |  |  |  |  |  |
| LR: Local Model | 0.731 (0.718 - 0.744) | 81.8% (79.7 - 83.7) | 49.7% (48.8 - 50.6) | 53.3% (52.5 - 54.2) | 17.1% (16.2 - 18.0) | 95.6% (95.0 - 96.1) | 0.282 |
| LR: Federated Global Model | 0.865 (0.855 - 0.876) | 78.2% (76.1 - 80.2) | 81.0% (80.3 - 81.7) | 80.7% (80.0 - 81.3) | 34.2% (32.6 - 35.8) | 96.7% (96.3 - 97.0) | 0.476 |
| LR: Federated Site-Personalised Model | 0.867 (0.856 - 0.878) | 74.2% (71.9 - 76.4) | 85.7% (85.0 - 86.3) | 84.4% (83.8 - 85.0) | 39.6% (37.8 - 41.4) | 96.3% (96.0 - 96.7) | 0.516 |
| DNN: Local Model | 0.622 (0.608 - 0.637) | 74.5% (72.3 - 76.7) | 43.8% (42.9 - 44.7) | 47.3% (46.4 - 48.1) | 14.4% (13.6 - 15.2) | 93.2% (92.5 - 93.8) | 0.241 |
| DNN: Federated Global Model | 0.876 (0.865 - 0.886) | 77.2% (74.9 - 79.2) | 82.3% (81.6 - 82.9) | 81.7% (81.0 - 82.3) | 35.5% (33.8 - 37.1) | 96.6% (96.2 - 96.9) | 0.486 |
| DNN: Federated Site-Personalised Model | 0.883 (0.873 - 0.893) | 78.2% (76.1 - 80.2) | 82.7% (82.0 - 83.4) | 82.2% (81.6 - 82.9) | 36.4% (34.7 - 38.1) | 96.8% (96.4 - 97.1) | 0.497 |
| **Bedfordshire Hospitals NHS Foundation Trust** |  |  |  |  |  |  |  |
| LR: Federated Global Model | 0.878 (0.851 - 0.904) | 83.4% (76.6 - 88.6) | 73.6% (70.8 - 76.2) | 74.8% (72.3 - 77.2) | 30.6% (26.3 - 35.3) | 97.0% (95.5 - 97.9) | 0.448 |
| DNN: Federated Global Model | 0.917 (0.893 - 0.942) | 89.7% (83.6 - 93.6) | 76.6% (73.9 - 79.1) | 78.2% (75.7 - 80.5) | 34.9% (30.2 - 39.8) | 98.1% (97.0 - 98.9) | 0.502 |

Tuning of the global models for individual sites, by performing an additional round of training on the local training set prior to prospective evaluation, led to a small improvement in DNN performance at PUH (with AUROC improving <0.01; DeLong p=0.0014) but not for OUH (DeLong p=0.262). For LR, site personalisation did not improve performance (DeLong p=0.269 & p=0.629 for PUH and OUH respectively; Table 3 & Figure 3). This finding suggests low levels of population distribution shifts for predictors between sites, and high generalisability of the global model.

### Explainable AI (XAI)

Coefficient analysis of the LR global model showed granulocyte counts, including Neutrophils and Eosinophils, Albumin and respiratory rate had the highest impact on model predictions. This finding is in keeping with results in previous work, and the recognised roles of these predictors in the inflammatory response^25, 26^. However, different to previous results, Haematocrit had a relatively larger coefficient possibly reflecting that coefficient analysis may identify co-variates/correlates. Shapley Additive exPlanations (SHAP) values, which provide a quantitate measure of the impact a feature has a models’ predictions, identified similar features as having greatest effect on the LR global model predictions. For the DNN, SHAP values showed that Eosinophil count has greatest impact on model predictions, similar to the findings with the previous XGBoost based CURIAL-Lab model^25^.

## Discussion

Best practice in health data research mandates risks of inadvertent or malicious disclosure should be mitigated as far as possible, irrespective of whether pseudonymisation is used^34–36^. Although AI methods have shown great promise for improving diagnostics, participation by healthcare providers who hold EHR data has been limited thereby reducing diversity of available training data and limiting potential healthcare improvements^11, 37–39^.

Here, we present a real-world deployment of an end-to-end FL pipeline in tandem with IoMT hardware in the UK’s NHS. We train, calibrate and validate a COVID-19 screening model for emergency care in a decentralised fashion, without centralising patient data from the four participating hospital groups, developing a user-friendly method for sites to participate without local technical expertise. Our micro-computing solution uses commercially available hardware and can be rapidly replicated across providers, achieving scalability at a low per-site cost (<£80). We propose that FL has potential to become a new standard-of-practice for privacy-prioritising health data research, reducing participation barriers and contributing to reduction of bias within training sets^7^, and allowing cross-border collaboration while maintaining data sovereignty.

While FL theoretically offers safeguards against leakage in case of interception attack, additional considerations are required to mitigate other security risks such as unauthorised access or code injection. The use of single-purpose client and server hardware reduced the risk of inadvertent trojan attack. We selected the most recent long-term support (LTS) release of Ubuntu (22.04.1 LTS), an enterprise Linux distribution with full commercial support, configured to automatically accept security updates. Clients were secured in line with local requirements, and participating sites were asked to physically safeguard devices following local processes for handing IT hardware containing pseudonymised data. Source code was available to participating sites for review. Data was held on the client in pseudonymised form for the period of analysis only, protected by the site network’s firewall, and switched off when not in use. Sites were directed to remove and destroy the microSD storage disk on completion of participation. Where required, firewall rules were instated by local IT/security teams to allow two-way traffic communication between the device and co-ordinating server via a single pre-agreed port. The co-ordinating server was hosted in a dedicated virtual machine on the Microsoft Azure platform [Redmond, California, USA], within an isolated virtual network and subject to the security considerations of the Azure platform ^40^. External communication was restricted to the pre-agreed port only, and the server was switched off when not in use for the present study. Messages between client and server contained only weights from within the trained model or summary results of evaluation, providing inherent protection against leakage if intercepted.

Our results find that federation provided significant performance improvement over training on a single-site (figure 2), bringing model performance in to a clinically-acceptable range (AUROC 0.917 during external validation; Table 3). Our findings show a more marked performance increase for deep neural networks, in keeping with other applications of federated deep learning^8, 17, 18, 41^. This possibly reflects the high data requirements of DNNs, which require large quantities of data to extract high-level features^42^, indicating that FL may be an enabling technology for deep-learning based medical AI^41^. When compared to the original XGBoost-based CURIAL-Lab model, our federated global DNN model achieves higher performance during the comparable external evaluation at BH (CURIAL-Lab AUROC 0.881, 95% CIs 0.851-0.912^25^, versus CURIAL-Fed-Lab AUROC 0.917, 0.893 - 0.942).

Strengths of our federated approach include the elimination of transfer of patient data off-premises as a prerequisite for participation, and without requiring use of a data-intermediary or Trusted Research Environment (TRE). We use the Raspberry Pi 4B device owing to commercial availability, high levels of support, and inexpensive removable storage medium (microSD; <£9 for 32Gb). To aid rapid deployment at scale, the microSD card of a configured Raspberry Pi can be imaged and cloned for new sites. Further, as the microSD cards are interchangeable, this approach may allow for a novel network of research devices to be maintained on providers’ premises, with new microSD cards sent to Trusts when deploying a new medical-AI application. Our study reports the largest number of patient encounters in a published secondary-care FL study to date, including routinely collected data from 160,000 presentations to acute and emergency services across three UK regions. As our pipeline is designed for the well-supported Ubuntu operating system, our solution may readily scale to hardware offering greater computational power for more demanding learning tasks or operate on existing hospital-owned hardware.

Notable limitations included that prior knowledge of the data format was required to allow harmonisation of feature names, unit values, and the representation of out-of-bounds values between sites. We approached this by providing Trusts with a data specification and dictionary, however future work may explore a role for complimentary PETs such as differential privacy or synthetic data where a more in-depth knowledge of the data sets is required^43, 44^. Further, future work would seek to implement a fully-autonomous data extraction pathway through direct EHR integration, although challenges of this are well described^45, 46^. The distributed nature of FL required that sensitivity and subgroup analyses are defined a priori, as only model weights and evaluation results are transmitted, potentially limiting researchers’ ability to rapidly investigate trends discovered within early results. FL, in combination with other PETs, do not create a trustless system and continue to require professional conduct during the manual stages of data and device handling. Lastly, the small physical footprint of micro-computing hardware may increase its susceptibility to loss or theft, requiring greater consideration towards physical security measures.

Our results show that, in this case, site-specific tuning did not significantly improve model performance, suggesting that levels of distribution shift between sites were small for the routinely collected data examined and confirming good generalisability of the global model. This finding may vary for different clinical scenarios, using clinical data where site-specific variations are more likely, for example, where they may be differences in sample preparation protocols between sites, or when working across international borders.

In conclusion, our work demonstrates an effective deployment of federated learning for the real-world emergency care setting. Future work may examine the extent to which increased diversity due to federation can improve model fairness, applications of similar techniques to different clinical questions, and methods to incentivise uptake by providers^41^.

## Supporting information

TRIPOD Checklist

## Data Availability

Data & Code Availability:
The code for our federated learning pipeline is available online alongside publication, via Github. Although, by design, patient data was not transferred within the FL for this study, AS, JY, DWE and DAC have previously had access to the raw data within a prior related evaluation study (Soltan et. al 2022). PD accessed and verified the data at BH. MAS & DRT accessed and verified the data at UHB. LDC accessed and verified the data at PUH. Data from OUH studied here are available from the Infections in Oxfordshire Research Database (https://oxfordbrc.nihr.ac.uk/research-themes-overview/antimicrobial-resistance-and-modernising-microbiology/infections-in-oxfordshire-research-database-iord/), subject to an application meeting the ethical and governance requirements of the Database. Data from UHB, PUH and BH are available on reasonable request to the respective trusts subject to NHS HRA requirements.

https://oxfordbrc.nihr.ac.uk/research-themes/modernising-medical-microbiology-and-big-infection-diagnostics/infections-in-oxfordshire-research-database-iord/

## Acknowledgments

We express our sincere thanks to all patients and staff across the four participating NHS trusts; Oxford University Hospitals, University Hospitals Birmingham, Bedfordshire Hospitals, and Portsmouth Hospitals University NHS Trusts.

## Declarations

DWE reports personal fees from Gilead, outside the submitted work; DAC reports personal fees from Oxford University Innovation, personal fees from BioBeats, personal fees from Sensyne Health, outside the submitted work. No other authors report any conflicts of interest.

## Contributions statement

AS conceived of and designed the study with design input from TZ, AT and DAC. AS wrote the federated learning code, developed the client/server hardware implementation, performed the analyses and wrote the manuscript. AT provided support with development of the FL setup. JY supported with earlier versions of code to harmonise data at two of the sites. DWE performed data extraction at OUH. PD (BH), LDC/AC (PUH), AS (OUH) and MAS/DT (UHB) operated the client devices at the respective sites. All authors reviewed and edited the manuscript.

## Data & Code Availability

The code for our federated learning pipeline is available online alongside publication, via Github. We additionally supply a flashable disk-image file representing the pre-configured environment loaded on to the Raspberry Pi 4B devices supplied to participating NHS trusts. Although, by design, patient data was not transferred within the FL for this study, AS, JY, DWE and DAC have previously had access to the raw data within a prior related evaluation study (Soltan et. al 2022^26^). PD accessed and verified the data at BH. MAS & DRT accessed and verified the data at UHB. LDC accessed and verified the data at PUH. Data from OUH studied here are available from the Infections in Oxfordshire Research Database (https://oxfordbrc.nihr.ac.uk/research-themes-overview/antimicrobial-resistance-and-modernising-microbiology/infections-in-oxfordshire-research-database-iord/), subject to an application meeting the ethical and governance requirements of the Database. Data from UHB, PUH and BH are available on reasonable request to the respective trusts subject to NHS HRA requirements.

## Supplementary Material

### Appendix A: Supplementary Methods

#### Description of Training & Evaluation Cohorts

OUH consists of four teaching hospitals, serving a population of 600 000 and providing tertiary referral services to the surrounding region. Data extraction considered all patients presenting to emergency and acute medical departments prior to the pandemic, between **December 1, 2018 and November 30, 2019**, and during first and second waves of the COVID-19 pandemic in the UK between **December 1, 2019 and March 6, 2021**. Confirmatory testing at OUH was by laboratory RT-PCR assay (Abbott Architect [Abbott, Maidenhead, UK], TaqPath [Thermo Fisher Scientific, Massachusetts, USA] and Public Health England-designed RNA-dependent RNA polymerase assays).

Training & evaluation cohorts at Portsmouth Hospitals University NHS Trust (PUH) considered all patients presenting to the Queen Alexandria Hospital (QAH), serving a population of 675,000 and offering tertiary referral services to the surrounding region. We included patients admitted prior to the pandemic between, between **January 01, 2019 and October 31, 2019**, and during first & second waves of the COVID-19 pandemic in the UK between **March 1, 2020 and February 28, 2021**. Confirmatory COVID-19 testing was by laboratory SARS-CoV-2 RT-PCR assay (Ct for positive result ≤36), considering any positive PCR result within 48hrs of admission as a true positive.

University Hospitals Birmingham NHS Foundation (UHB) Trust participated in federated training & testing only. Data extraction for training & testing considered all patients admitted to The Queen Elizabeth Hospital, Birmingham, prior to the pandemic between **December 01, 2018 and November 30, 2019**, and during the first wave of the COVID-19 pandemic between **December 01, 2019 and October 29, 2020**. The Queen Elizabeth Hospital is a large tertiary referral unit within the UHB group which provides healthcare services for a population of 2.2 million across the West Midlands. Confirmatory COVID-19 testing was performed by laboratory SARS-CoV-2 RT-PCR assay (Ct for positive result ≤36).

Bedfordshire Hospitals NHS Foundation Trust (BHT) participated in federated evaluation only. Data extraction for the evaluation considered all patients admitted to Bedford Hospital between **January 1, 2021 and March 31, 2021**. BHT provides healthcare services for a population of around 620,000 in Bedfordshire. Confirmatory COVID-19 testing was performed on the day of admission by point-of-care PCR based nucleic acid testing [SAMBA-II & Panther Fusion System, Diagnostics in the Real World, UK, and Hologic, USA]. The Ct for a positive clinical result was ≤36. In an evaluation of the SAMBA-II against laboratory RT-PCR testing, the SAMBA-II achieved sensitivity of 96.9% and specificity of 99.1%^47, 48^.

We report sensitivity, specificity, positive and negative predictive values (PPV and NPV), AUROC and F1 alongside 95% CIs, comparing model predictions to results of confirmatory viral testing (laboratory PCR and SAMBA-II). 95% Confidence intervals for sensitivity, specificity and predictive values were computed using Wilson’s Method^49^, and for AUROC with DeLong’s method^33^.

### Data Extraction

For PUH, UHB and BH, data extraction and pseudonymisation was performed by a member of staff employed by the respective trust within the participating NHS trust’s premises. Data extraction for OUH was performed within the shared research network that exists between the NHS trust and Oxford University (Oxford NIHR Biomedical Research Campus) via the Big Data Institute. The pseudonomised data was extracted from electronic health records according to the pre-provided extraction criteria as comma-delimited files (CSVs; Supplementary Table S1 & Methods) and transferred to a specified folder on the Raspberry Pi using locally-approved USB flash drives or secure file transfer protocol (sFTP) within the local network.

### The FedAvg Algorithm

Model updates were performed using the FedAvg algorithm, in which the server collects weights within locally trained models and updates the global model by calculating a weighted average^10^. The weight attributed to model updates from each client is weighted by the number of samples contributed to the training process.

### Client-Server Communication

We implemented client-server federated learning using the Flower library (v 1.2.0). Clients communicated with the server using in-built protocols via a dedicated port. Where required, firewall rules were implemented by local IT/security teams to allow communication between the client device and the server through the pre-agreed port. Media Access Control (MAC) addresses were made available where required to facilitate network controls. Messages between client and server contained only weights from within the trained model or summary results of evaluation. Data was held on the Raspberry Pi in pseudonymised form and was not transferred between client and server.

### Device Security

To ensure security of the Raspberry Pi 4B devices, we selected the Ubuntu Desktop OS owing to its commercially security support. We selected the most up to date available version of Ubuntu at the time of deployment (22.04.1 LTS), configuring the devices to automatically install security patches. Devices were password protected, and participating sites were asked to physically safeguard the devices following local processes for handling of IT hardware. We made source code of the pipeline available to participating sites. On study completion, sites were asked to destroy the microSD storage card. Devices were switched off when not in use. The federated server was hosted within a dedicated virtual machine on the Microsoft Azure platform, as a Standard D2s v3 machine with 8 Gb RAM, and running the Ubuntu Server operating system. A virtual network and associated security group was implemented, permitting communication with the server only through the approved port. Both server and clients were switched off when not in use.

### Software Libraries & Dependencies

Ubuntu Desktop & Server 22.04.01 LTS Flower v 1.2.0

Pandas 1.5.0

Tensorflow & Keras 2.10.0 Scikit-learn 1.1.2

SciPy 1.9.1

Numpy 1.23.3

Statsmodels 0.13.2

SHAP 0.41.0

**Supplementary Table S1:** Clinical data fields, extracted from training and prospectively/externally validating NHS sites, for all patients admitted to the trusts during the study periods. (Table reproduced from Soltan et. al. 2022^26^). Premorbid clinical data were not analysed within this study.

| Clinical Descriptors: | Presentation Blood Tests: | Presentation Blood Gas: | Premorbid Clinical Data |
| --- | --- | --- | --- |
| Study ID | PresentationHAEMOGLOBIN | PresentationPOCT pC02 | BaselineHAEMOGLOBIN |
| Presentation Date | PresentationWHITE CELLS | PresentationPOCT sO2 | BaselineWHITE CELLS |
| Ethnicity | PresentationPLATELETS | PresentationPOCT pO2 | BaselinePLATELETS |
| Age at presentation | PresentationMEAN CELL VOL. | PresentationPCT cBASE(Ecf) | BaselineMEAN CELL VOL. |
| Gender (M/F) | PresentationRED CELL COUNT | PresentationPCT CO3(P,st) | BaselineRED CELL COUNT |
| Comorbidities (ICD10) | PresentationNEUTROPHILS | PresentationPOCT Hctc | BaselineNEUTROPHILS |
| Outcome | PresentationHAEMATOCRIT | PresentationPOCT FO2Hb | BaselineHAEMATOCRIT |
| **Vital Signs:** | PresentationLYMPHOCYTES | PresentationPOCT ctO2c | BaselineLYMPHOCYTES |
| AdmissionRespRate | PresentationMEAN CELL HGB | PresentationPOCT cGLU | BaselineMEAN CELL HGB |
| AdmissionHeartRate | PresentationMONOCYTES | PresentationPOCT cK+ | BaselineMONOCYTES |
| AdmissionBloodPressure | PresentationEOSINOPHILS | PresentationPOCT cNA+ | BaselineEOSINOPHILS |
| AdmissionSpO2 | PresentationBASOPHILS | PresentationPOCT cLAC | BaselineBASOPHILS |
| AdmissionOxygenDeliveryDevice | Presentation MCH | PresentationPOCT cCA++ | BaselineMEAN CELL HGB CONC |
| AdmissionTemperature | PresentationMPV |  | BaselineSODIUM |
| **Microbiology:** | PresentationNRBC A |  | BaselineALBUMIN |
| SARS-CoV-2 PCR | PresentationNRBC % |  | BaselineALK.PHOSPHATASE |
| SARS-CoV-2 RESULT TYPE | PresentationSODIUM |  | BaselineALT |
| SARS-CoV-2 Antigen Test Result | PresentationALBUMIN |  | BaselineUREA |
| INFLUENZAPCR | PresentationALK.PHOSPHATASE |  | BaselineBILIRUBIN |
| RespiratoryPCR (Biofire) | PresentationALT |  | BaselineCREATININE |
|  | PresentationUREA |  | BaselineeGFR |
|  | PresentationBILIRUBIN |  | BaselinePOTASSIUM |
|  | PresentationCREATININE |  | BaselineCALCIUM |
|  | PresentationeGFR |  | BaselineADJUSTED CALC. |
|  | PresentationPOTASSIUM |  | BaselineCRP |
|  | PresentationCALCIUM |  | BaselineProthromb. Time |
|  | PresentationADJUSTED CALC. |  | BaselineAPTT |
|  | PresentationPHOSPHATE |  | BaselineINR |
|  | PresentationCRP |  | BaselinePOCT pC02 |
|  | PresentationProthromb. Time |  | BaselinePOCT sO2 |
|  | PresentationPOCT ctHb |  | BaselinePOCT pO2 |
|  | PresentationGLUCOSE |  | BaselinePCT cBASE(Ecf)c |
|  | PresentationAPTT |  | BaselinePCT CO3(P,st)c |
|  | PresentationINR |  | BaselinePOCT Hctc |
|  |  |  | BaselinePOCT FO2Hb |
|  |  |  | BaselinePOCT ctO2c |
|  |  |  | BaselinePOCT Cglu |
|  |  |  | BaselinePOCT cK+ |
|  |  |  | BaselinePOCT cNA+ |
|  |  |  | BaselinePOCT cLAC |
|  |  |  | BaselinePOCT cCA++ |

**Supplementary Figure S1:**
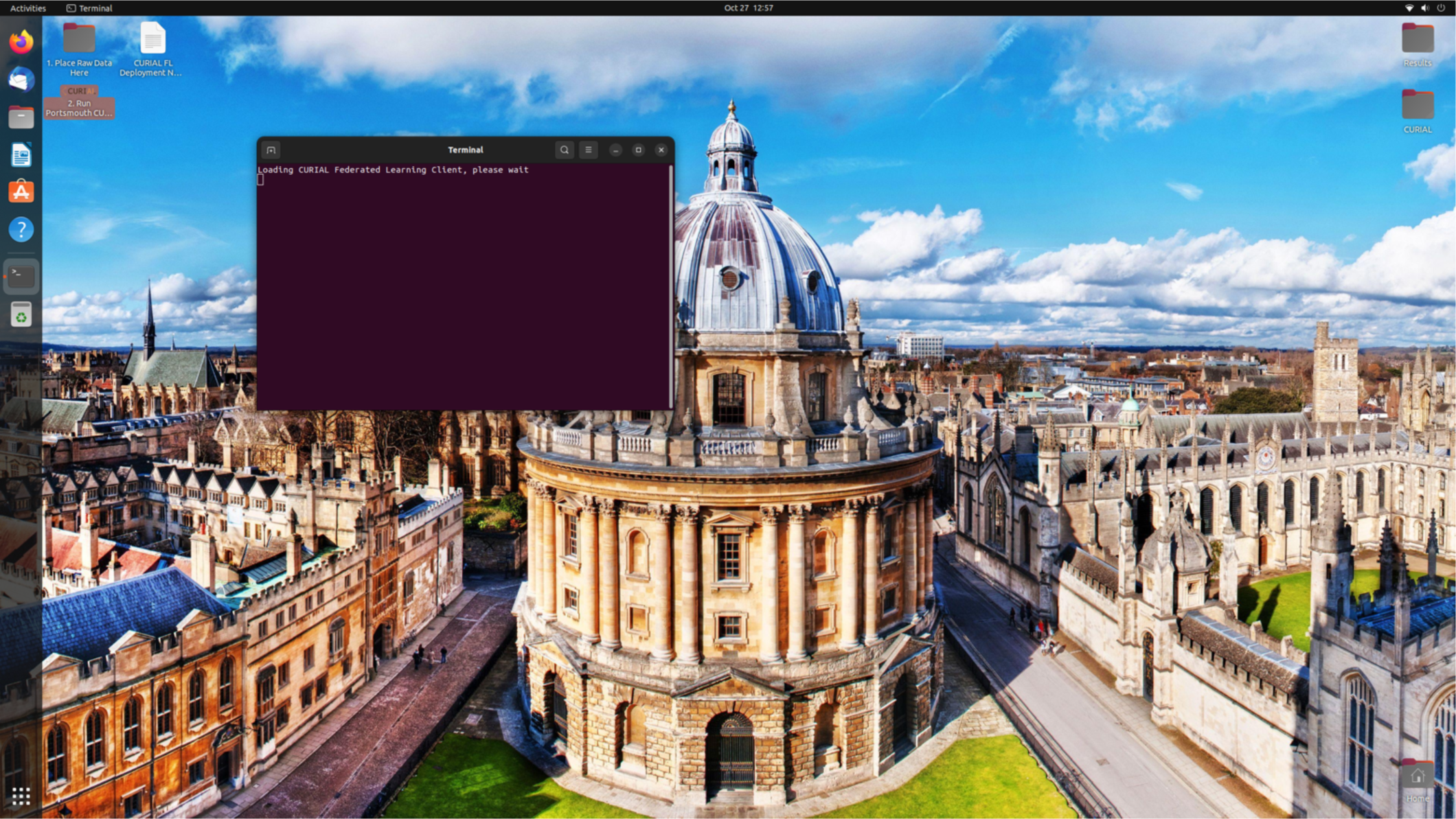
User interface for participating NHS Trusts on loading the Raspberry Pi 4B devices, showing the two-step participation process. Participating sites were requested to upload data files into a pre-agreed folder (‘1 Place Raw Data Here’) as Comma-Separated Value files, and select option ‘2 Run Client’ to operate the FL pipeline.

## Appendix B: Supplementary Results

### Cohort summaries

**Supplementary Table S2:**
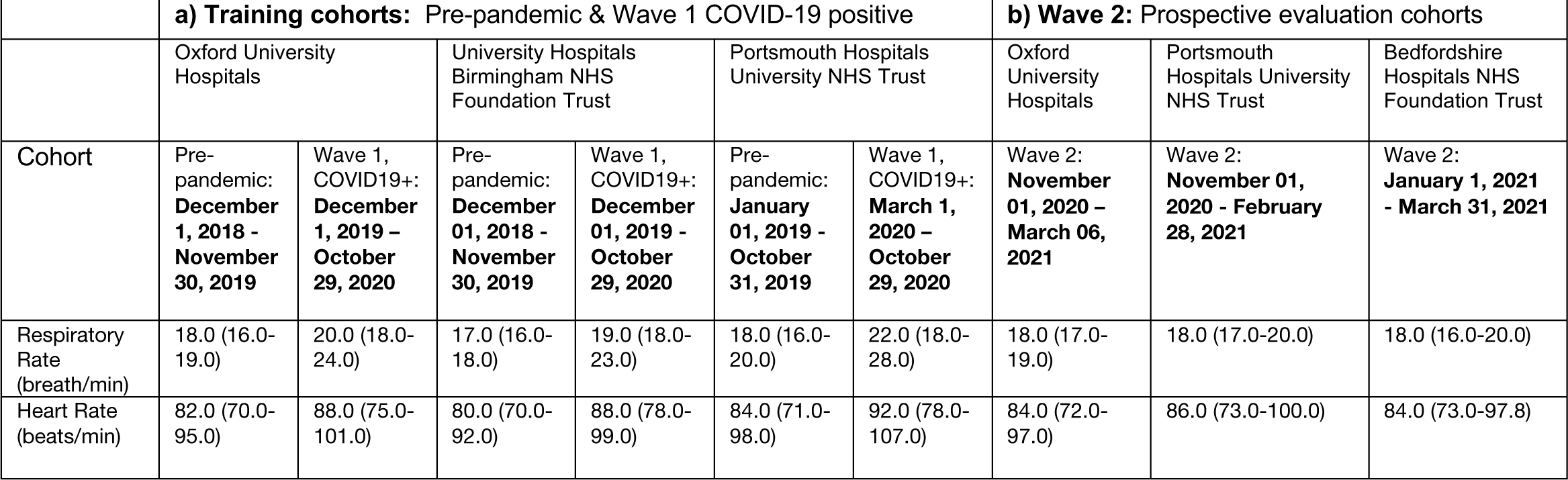
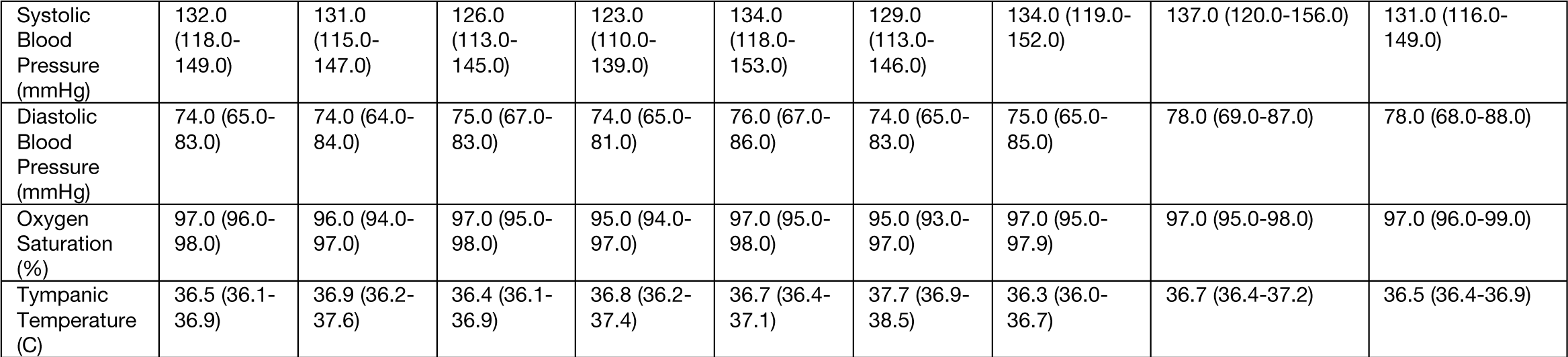
Distribution of vital signs, reported as median and interquartile ranges, for each patient cohort.

|  | a) Training cohorts: Pre-pandemic & Wave 1 COVID-19 positive |  |  |  |  |  | b) Wave 2: Prospective evaluation cohorts |  |  |
| --- | --- | --- | --- | --- | --- | --- | --- | --- | --- |
|  | Oxford University Hospitals |  | University Hospitals Birmingham NHS Foundation Trust |  | Portsmouth Hospitals University NHS Trust |  | Oxford University Hospitals | Portsmouth Hospitals University NHS Trust | Bedfordshire Hospitals NHS Foundation Trust |
| Cohort | Pre-pandemic: December 1, 2018 - November 30, 2019 | Wave 1, COVID19+: December 1, 2019 - October 29, 2020 | Pre-pandemic: December 01, 2018 - November 30, 2019 | Wave 1, COVID19+: December 01, 2019 - October 29, 2020 | Pre-pandemic: January 01, 2019 - October 31, 2019 | Wave 1, COVID19+: March 1, 2020 - October 29, 2020 | Wave 2: November 01, 2020 - March 06, 2021 | Wave 2: November 01, 2020 - February 28, 2021 | Wave 2: January 1, 2021 - March 31, 2021 |
| Respiratory Rate (breath/min) | 18.0 (16.0-19.0) | 20.0 (18.0-24.0) | 17.0 (16.0-18.0) | 19.0 (18.0-23.0) | 18.0 (16.0-20.0) | 22.0 (18.0-28.0) | 18.0 (17.0-19.0) | 18.0 (17.0-20.0) | 18.0 (16.0-20.0) |
| Heart Rate (beats/min) | 82.0 (70.0-95.0) | 88.0 (75.0-101.0) | 80.0 (70.0-92.0) | 88.0 (78.0-99.0) | 84.0 (71.0-98.0) | 92.0 (78.0-107.0) | 84.0 (72.0-97.0) | 86.0 (73.0-100.0) | 84.0 (73.0-97.8) |
| Systolic Blood Pressure (mmHg) | 132.0 (118.0-149.0) | 131.0 (115.0-147.0) | 126.0 (113.0-145.0) | 123.0 (110.0-139.0) | 134.0 (118.0-153.0) | 129.0 (113.0-146.0) | 134.0 (119.0-152.0) | 137.0 (120.0-156.0) | 131.0 (116.0-149.0) |
| Diastolic Blood Pressure (mmHg) | 74.0 (65.0-83.0) | 74.0 (64.0-84.0) | 75.0 (67.0-83.0) | 74.0 (65.0-81.0) | 76.0 (67.0-86.0) | 74.0 (65.0-83.0) | 75.0 (65.0-85.0) | 78.0 (69.0-87.0) | 78.0 (68.0-88.0) |
| Oxygen Saturation (%) | 97.0 (96.0-98.0) | 96.0 (94.0-97.0) | 97.0 (95.0-98.0) | 95.0 (94.0-97.0) | 97.0 (95.0-98.0) | 95.0 (93.0-97.0) | 97.0 (95.0-97.9) | 97.0 (95.0-98.0) | 97.0 (96.0-99.0) |
| Tympanic Temperature (C) | 36.5 (36.1-36.9) | 36.9 (36.2-37.6) | 36.4 (36.1-36.9) | 36.8 (36.2-37.4) | 36.7 (36.4-37.1) | 37.7 (36.9-38.5) | 36.3 (36.0-36.7) | 36.7 (36.4-37.2) | 36.5 (36.4-36.9) |

**Supplementary Table S3:** Distribution of blood test features, reported as median and interquartile ranges, for each patient cohort.

|  | a) Training cohorts: Pre-pandemic & Wave 1 COVID-19 positive |  |  |  |  |  | b) Wave 2: Prospective evaluation cohorts |  |  |
| --- | --- | --- | --- | --- | --- | --- | --- | --- | --- |
|  | Oxford University Hospitals |  | University Hospitals Birmingham NHS Foundation Trust |  | Portsmouth Hospitals University NHS Trust |  | Oxford University Hospitals | Portsmouth Hospitals University NHS Trust | Bedfordshire Hospitals NHS Foundation Trust |
| Cohort | Pre-pandemic: December 1, 2018 - November 30, 2019 | Wave 1, COVID19+: December 1, 2019 – October 29, 2020 | Pre-pandemic: December 01, 2018 - November 30, 2019 | Wave 1, COVID19+: December 01, 2019 - October 29, 2020 | Pre-pandemic: January 01, 2019 - October 31, 2019 | Wave 1, COVID19+: March 1, 2020 – October 29, 2020 | Wave 2: November 01, 2020 – March 06, 2021 | Wave 2: November 01, 2020 - February 28, 2021 | Wave 2: January 1, 2021 - March 31, 2021 |
| HAEMOGLOBIN (g/L) | 130.0 (116.0-142.0) | 131.0 (115.0-143.0) | 131.0 (116.0-143.0) | 128.5 (114.0-141.0) | 127.0 (113.0-139.0) | 128.0 (113.0-142.0) | 130.0 (114.0-143.0) | 127.0 (113.0-140.0) | 134.0 (119.0-146.0) |
| WHITE CELLS (10<sup>9</sup> l<sup>-1</sup>) | 8.52 (6.57-11.24) | 6.74 (5.04-9.46) | 8.6 (6.7-11.2) | 7.15 (5.5-10.1) | 9.6 (7.3-12.7) | 7.0 (5.0-9.8) | 8.89 (6.65-12.01) | 9.3 (7.0-12.4) | 9.2 (6.95-12.5) |
| PLATELETS (10<sup>9</sup> l<sup>-1</sup>) | 248.0 (199.0-306.0) | 214.0 (160.0-282.5) | 237.0 (188.5-292.5) | 214.5 (162.0-291.75) | 245.0 (196.0-304.0) | 203.0 (157.0-255.0) | 250.0 (198.0-313.0) | 246.0 (194.0-309.0) | 246.0 (196.25-310.0) |
| MEAN CELL VOL (fl) | 90.1 (86.5-93.9) | 90.1 (86.3-94.1) | 88.6 (84.4-92.6) | 87.35 (83.28-91.9) | 86.6 (82.25-92.67) | 88.8 (85.2-92.2) | 90.2 (86.6-94.2) | 90.0 (86.3-93.5) | 88.0 (85.0-92.0) |
| NEUTROPHILS (10<sup>9</sup> l<sup>-1</sup>) | 5.83 (4.11-8.5) | 4.93 (3.34-7.28) | 5.8 (4.2-8.4) | 5.5 (3.9-8.0) | 7.1 (4.9-10.2) | 5.3 (3.6-8.0) | 6.4 (4.37-9.48) | 6.8 (4.7-9.8) | 6.8 (4.72-9.72) |
| HAEMATOCRIT | 0.39 (0.35-0.42) | 0.4 (0.36-0.43) | 0.39 (0.35-0.42) | 0.39 (0.35-0.42) | 0.38 (0.34-0.42) | 0.38 (0.34-0.42) | 0.39 (0.35-0.43) | 0.38 (0.34-0.42) | 0.39 (0.35-0.43) |
| LYMPHOCYTES (10<sup>9</sup> l<sup>-1</sup>) | 1.46 (0.96-2.07) | 0.98 (0.65-1.39) | 1.5 (1.0-2.1) | 0.95 (0.62-1.4) | 1.4 (0.9-2.0) | 0.8 (0.5-1.2) | 1.3 (0.85-1.88) | 1.3 (0.8-1.9) | 1.27 (0.85-1.83) |
| MONOCYTES (10<sup>9</sup> l<sup>-1</sup>) | 0.65 (0.49-0.87) | 0.48 (0.34-0.72) | 0.6 (0.5-0.9) | 0.45 (0.3-0.67) | 0.7 (0.5-0.9) | 0.5 (0.4-0.8) | 0.65 (0.47-0.89) | 0.7 (0.5-0.9) | 0.67 (0.48-0.92) |
| EOSINOPHILS (10<sup>9</sup> l<sup>-1</sup>) | 0.1 (0.04-0.2) | 0.01 (0.0-0.06) | 0.1 (0.0-0.2) | 0.01 (0.0-0.04) | 0.1 (0.0-0.2) | 0.0 (0.0-0.0) | 0.07 (0.02-0.16) | 0.1 (0.0-0.1) | 0.06 (0.02-0.16) |
| BASOPHILS (10<sup>9</sup> l<sup>-1</sup>) | 0.04 (0.03-0.06) | 0.02 (0.01-0.03) | 0.0 (0.0-0.1) | 0.02 (0.01-0.03) | 0.0 (0.0-0.1) | 0.0 (0.0-0.1) | 0.04 (0.02-0.06) | 0.0 (0.0-0.1) | 0.05 (0.03-0.07) |
| SODIUM (mM) | 138.0 (136.0-140.0) | 136.0 (133.0-139.0) | 138.0 (135.0-140.0) | 138.0 (135.0-140.0) | 137.0 (134.0-139.0) | 136.0 (133.0-139.0) | 138.0 (135.0-140.0) | 137.0 (134.0-139.0) | 138.0 (136.0-140.0) |
| ALBUMIN (g/L) | 36.0 (32.0-39.0) | 32.0 (28.0-36.0) | 39.0 (34.0-43.0) | 30.0 (26.0-33.0) | 36.0 (32.0-40.0) | 33.0 (29.0-36.0) | 36.0 (31.0-39.0) | 36.0 (32.0-39.0) | 35.0 (31.0-39.0) |
| ALKALINE PHOSPHATASE (IU/L) | 81.0 (65.0-106.0) | 80.0 (63.0-106.0) | 82.0 (66.0-106.0) | 76.0 (59.25-101.0) | 85.0 (67.0-114.0) | 88.0 (67.0-116.5) | 84.0 (66.0-112.0) | 89.0 (71.0-119.0) | 94.0 (74.0-122.0) |
| ALT (IU/L) | 18.0 (13.0-28.0) | 25.0 (16.25-40.0) | 18.0 (12.0-28.0) | 27.0 (17.0-44.0) | 18.0 (13.0-29.0) | 24.0 (16.0-39.0) | 20.0 (14.0-33.0) | 19.0 (13.0-31.0) | 20.0 (13.0-31.0) |
| UREA (mM) | 5.4 (4.1-7.7) | 5.8 (4.1-8.8) | 5.2 (3.8-7.4) | 6.1 (4.0-10.1) | 6.0 (4.3-8.8) | 8.0 (5.4-12.1) | 5.7 (4.2-8.4) | 6.2 (4.5-9.1) | 5.8 (4.2-8.3) |
| BILIRUBIN (umol/L) | 9.0 (6.0-13.0) | 9.0 (6.0-13.0) | 9.0 (6.0-14.0) | 11.0 (8.0-15.0) | 10.0 (7.0-15.0) | 10.0 (8.0-14.0) | 9.0 (6.0-14.0) | 10.0 (7.0-15.0) | 10.0 (7.0-14.0) |
| CREATININE (umol/L) | 74.0 (60.0-95.0) | 79.0 (64.0-105.0) | 76.0 (61.0-96.0) | 82.0 (64.0-111.0) | 75.0 (60.0-101.0) | 90.0 (71.0-129.0) | 74.0 (60.0-97.0) | 78.0 (62.0-104.0) | 81.0 (65.25-104.0) |
| eGFR (ml/min) | 83.0 (60.0-150.0) | 79.0 (54.0-150.0) | 82.0 (59.0-90.0) | 75.0 (49.75-90.0) | 79.0 (55.0-90.0) | 63.0 (43.0-86.0) | 84.0 (58.0-150.0) | 76.0 (52.0-90.0) | 75.5 (54.0-90.0) |
| POTASSIUM (mM) | 4.0 (3.7-4.3) | 4.0 (3.7-4.3) | 4.2 (3.9-4.5) | 4.1 (3.7-4.5) | 4.1 (3.8-4.4) | 4.0 (3.7-4.3) | 4.0 (3.8-4.4) | 4.1 (3.8-4.4) | 4.3 (4.0-4.6) |
| CRP (mg/L) | 9.8 (2.5-44.8) | 64.95 (19.7-139.48) | 11.0 (3.0-57.0) | 102.0 (45.0-175.0) | 14.0 (4.0-60.0) | 81.0 (27.75-159.25) | 16.7 (3.5-69.2) | 14.0 (4.0-70.0) | 10.85 (2.8-49.05) |

### Data-completeness summaries

**Supplementary Table S5:** Numbers of participants with data-completeness for each predictor, across each evaluation cohort.

|  | **Wave 2: Prospective evaluation cohorts** |  |  |
| --- | --- | --- | --- |
|  | Oxford University Hospitals | Portsmouth Hospitals University NHS Trust | Bedfordshire Hospitals NHS Foundation Trust |
| Cohort | Wave 2:<br>**November 01, 2020 – March 06, 2021** | Wave 2:<br>**November 01, 2020 - February 28, 2021** | Wave 2:<br>**January 1, 2021 - March 31, 2021** |
| HAEMOGLOBIN (g/L) | 18275/18543 (98.6%) | 13210/13260 (99.6%) | 1183/1183 (100.0%) |
| WHITE CELLS (10<sup>9</sup> l<sup>-1</sup>) | 18275/18543 (98.6%) | 13208/13260 (99.6%) | 1183/1183 (100.0%) |
| PLATELETS (10<sup>9</sup> l<sup>-1</sup>) | 18262/18543 (98.5%) | 13191/13260 (99.5%) | 1178/1183 (99.6%) |
| MEAN CELL VOL (fl) | 18275/18543 (98.6%) | 13204/13260 (99.6%) | 1183/1183 (100.0%) |
| NEUTROPHILS (10<sup>9</sup> l<sup>-1</sup>) | 18174/18543 (98.0%) | 13202/13260 (99.6%) | 1183/1183 (100.0%) |
| HAEMATOCRIT | 18275/18543 (98.6%) | 13208/13260 (99.6%) | 1183/1183 (100.0%) |
| LYMPHOCYTES (10<sup>9</sup> l<sup>-1</sup>) | 18187/18543 (98.1%) | 13202/13260 (99.6%) | 1183/1183 (100.0%) |
| MONOCYTES (10<sup>9</sup> l<sup>-1</sup>) | 18209/18543 (98.2%) | 13205/13260 (99.6%) | 1183/1183 (100.0%) |
| EOSINOPHILS (10<sup>9</sup> l<sup>-1</sup>) | 18209/18543 (98.2%) | 13202/13260 (99.6%) | 1183/1183 (100.0%) |
| BASOPHILS (10<sup>9</sup> l<sup>-1</sup>) | 18205/18543 (98.2%) | 13205/13260 (99.6%) | 1183/1183 (100.0%) |
| SODIUM (mM) | 18206/18543 (98.2%) | 12700/13260 (95.8%) | 1179/1183 (99.7%) |
| ALBUMIN (g/L) | 16298/18543 (87.9%) | 12431/13260 (93.7%) | 1166/1183 (98.6%) |
| ALKALINE PHOSPHATASE (IU/L) | 16199/18543 (87.4%) | 12431/13260 (93.7%) | 1117/1183 (94.4%) |
| ALT (IU/L) | 16036/18543 (86.5%) | 12411/13260 (93.6%) | 1042/1183 (88.1%) |
| UREA (mM) | 18171/18543 (98.0%) | 12693/13260 (95.7%) | 1147/1183 (97.0%) |
| BILIRUBIN (umol/L) | 16050/18543 (86.6%) | 12412/13260 (93.6%) | 944/1183 (79.8%) |
| CREATININE (umol/L) | 18216/18543 (98.2%) | 12703/13260 (95.8%) | 1178/1183 (99.6%) |
| eGFR (ml/min) | 18171/18543 (98.0%) | 12703/13260 (95.8%) | 1178/1183 (99.6%) |
| POTASSIUM (mM) | 17870/18543 (96.4%) | 12154/13260 (91.7%) | 1062/1183 (89.8%) |
| CRP (mg/L) | 15506/18543 (83.6%) | 12274/13260 (92.6%) | 1140/1183 (96.4%) |
| Respiratory Rate (breath/min) | 18486/18543 (99.7%) | 11715/13260 (88.3%) | 1183/1183 (100.0%) |
| Heart Rate (beats/min) | 18531/18543 (99.9%) | 11716/13260 (88.4%) | 1182/1183 (99.9%) |
| Systolic Blood Pressure (mmHg) | 18530/18543 (99.9%) | 11715/13260 (88.3%) | 1177/1183 (99.5%) |
| Diastolic Blood Pressure (mmHg) | 18529/18543 (99.9%) | 11715/13260 (88.3%) | 1177/1183 (99.5%) |
| Oxygen Saturation (%) | 18524/18543 (99.9%) | 11716/13260 (88.4%) | 1183/1183 (100.0%) |
| Tympanic Temperature (C) | 18469/18543 (99.6%) | 11714/13260 (88.3%) | 1183/1183 (100.0%) |
| Oxygen Delivery Device | 18462/18543 (99.6%) | 11722/13260 (88.4%) | 1183/1183 (100.0%) |

**Supplementary Figure S2:**
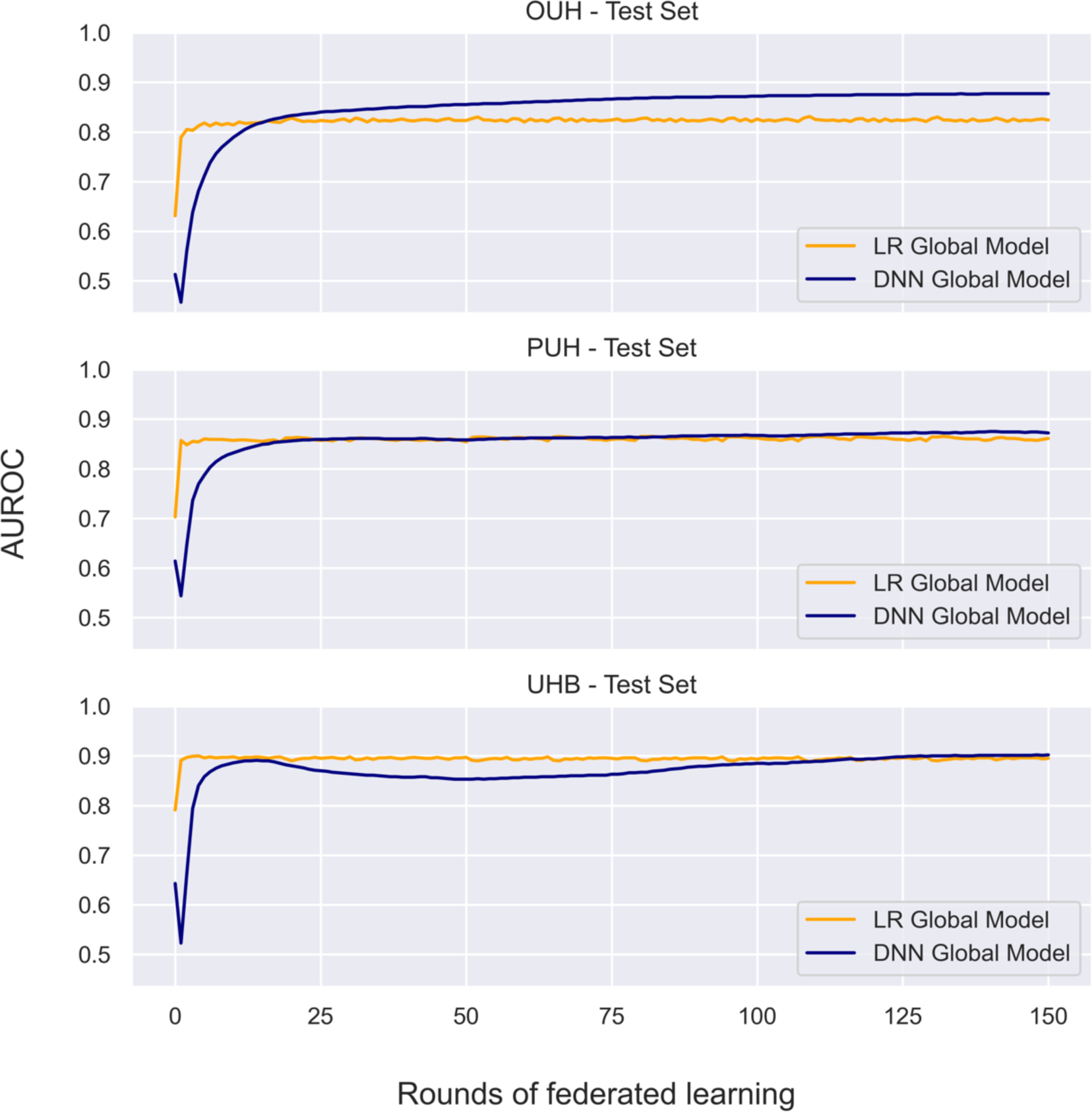
Curves showing iterative improvement in performance (AUROC) of the global model after each round of federated training, evaluated on the held-out test set for each site participating in training.

